# Randomised trial of not providing booster diphtheria-tetanus-pertussis (DTP) vaccination after measles vaccination and child survival: A failed trial

**DOI:** 10.1101/2025.10.29.25339081

**Authors:** Jane Agergaard, Sebastian Nielsen, Christine S. Benn, Peter Aaby

**Author notes:** Corresponding author: Peter Aaby.

## Abstract

**Background:** In low-income settings, WHO recommended a booster-dose of DTP (DTP4) and OPV (OPV4) at 18 months of age. Previous studies found DTP after MV to be associated with higher female mortality. We conducted a randomised trial (RCTs) in Guinea-Bissau, in which children from 18 months of age were randomised to receive DTP4+OPV4 vs OPV4-only to assess the impact on survival and hospitalisations, overall and by sex.

**Methods:** The trial was conducted from 2005-2012. Children were randomised and followed from 18 months to 4 years of age. Hospitalisations were captured through a surveillance system at the main paediatric ward, and deaths were detected through a demographic surveillance system in the study area and through yearly home visits to the study participants. Data was analysed in Cox proportional hazards models with age as underlying time scale. The RCT aimed to enrol 6,000 children based on an expected annual mortality rate of 3%. During the trial period many interventions, including many national health campaigns, were carried out.

**Results:** The trial enrolled 5,960 children of which 95% (5,674) were included in the analysis. Annual non-accidental mortality rate was 0.55%, 82% lower than the expected rate. There was no difference in mortality between the randomisation groups. With 30 and 35 deaths, the hazard ratio (HR) for DTP4+OPV4 vs OPV4-only was 0.84 (95% CI=0.52-1.37); 0.86 (0.42-1.76) in males and 0.82 (0.42-1.60) in females. The HR for hospitalisations was 0.91 (0.77-1.07); 0.87 (0.70-1.09) in males; 0.96 (0.75-1.22) in females).

**Conclusion:** The study found no negative effect of DTP4 and no sex-differential effect of DTP4, but it was strongly underpowered. Furthermore, due to the large number of health interventions, not envisioned at the initiation of the trial, a limited part of the follow-up was a comparison between DTP4+OPV4 vs OPV4 as the most recent vaccinations.

## INTRODUCTION

During the 1980s and 1990s, several studies suggested that measles vaccine (MV) had effects on mortality which could not be explained by prevention of measles infection. Standard-titre-measles-vaccine (STMV) reduced mortality much more than expected; e.g. after the introduction of MV the general mortality level in the affected age group declined more than 50% (1,2). Several studies indicated that these beneficial non-specific effects (NSEs) were more pronounced for females than males (3,4).

On the other hand, high-titre-measles-vaccine (HTMV) was protective against measles infection, but surprisingly, it was associated with higher female mortality, when tested against STMV (5,6). Hence, NSEs could be beneficial or deleterious and they were often sex-differential.

With 9 months being the WHO-recommended age of MV, children could not be randomised to not receive MV at that age. The first RCT of STMV to explore the NSEs was therefore designed as a two-dose trial, in which the children received STMV or inactivated polio vaccine (IPV) at 6 months of age and then all children received the recommended dose of MV at 9 months age. There was no beneficial effect of early STMV (7,8). *Post hoc* analyses revealed that many children had not received all three recommended doses of DTP before being enrolled in the MV trial and therefore received DTP with or after the first dose of MV (8). Among such children, there was a negative effect of receiving DTP after MV, which was strongest for females. This surprising observation provided a resolution to the enigma of why the HTMV was associated with higher female mortality: HTMV had been given so early at 4-5 months of age that most children had not received all doses of DTP and therefore received DTP after HTMV (5). Reanalysis of the original RCTs confirmed that if DTP was not administered after HTMV, HTMV did not have a negative effect. Hence, DTP after MV could have negative NSEs, particularly for females.

Subsequent studies supported a negative effect of receiving DTP after MV and other studies revealed that also DTP with MV could be less beneficial than MV alone (Supplementary Table 1, 9-14).

As a result, a randomised trial was designed to test the optimal policy for children, who were coming to the health centre to receive *either* DTP <u>with</u> MV *or* DTP <u>after</u> MV. Children due to receive MV were randomised to receive DTP+OPV or OPV with MV (arms 1+3, Supplementary Figure 1). Not many children were missing both MV and DTP and this part of the trial was stopped and reported in 2011 (15).

Children, who had already received MV, were randomised to DTP+OPV or OPV (arms 2+4, Supplementary Figure 1). These children were almost exclusively children who had received the primary series of 3 DTP vaccines in infancy and came to receive the DTP-booster (DTP4) vaccine at 18 months of age (arm 4, Supplementary Figure 1). In the present paper, we present the results of this part of the trial.

## METHODS

### Trial design

The trial was a community-based RCT randomising children at individual level to receive DTP4 (DTP-booster) with OPV4 vs OPV4-only between 18 months and 4 years of age. The children had already received the primary series of DTP recommended at 6, 10 and 14 weeks of age and the MV recommended at 9 months of age.

The main objectives were to assess whether not providing DTP4 would be associated with lower child mortality and less severe morbidity (hospitalisations) in a low-income country with a high pressure of infections and whether there were sex-differences in these outcomes.

### Participants

The RCT was initiated in October 2005 at the Bandim Health Project (BHP) in Guinea-Bissau (www.bandim.org), a Health and Demographic Surveillance System (HDSS) site, which covers six districts with approx. 100,000 inhabitants of the capital Bissau. Within this area, 3,500 children were born each year. BHP assistants visit all 6,000 houses monthly to register new pregnancies and new births. Once a birth is detected, a form is filled in and major background socio-economic risk factors (maternal education, electricity in the household, number of children etc.) are registered. Around 60-70% of children were delivered at two maternities, and these births were registered daily. Vaccinations are provided and registered at three health centres in the study area.

Every three months, field assistants visit all children under three years of age and data are collected on breastfeeding, hospitalisations, mid-upper-arm-circumference (MUAC), vaccinations, infections, movements, and survival.

Children were eligible for inclusion into the trial if they were 18 months old, had no overt illness, and had received the three primary doses of DTP vaccine and MV.

The enrolment team consisted of a study physician, nurses, and field workers. The team worked at the three health centres in the study area on different weekdays. Mothers of eligible children were invited to come to the health centre to take part in a trial. Here, the mothers received an oral explanation in Portuguese Creole and a written explanation in Portuguese from the physician. Participation was voluntary. Mothers/guardians were asked to provide oral and written consent. The physician performed a medical examination of whether the child was healthy enough to be included in a trial.

### Randomisation and masking

Eligible participants were randomised 1:1 to receive DTP4+OPV4 (standard of care at the initiation of the trial) or OPV4. The randomisation lots were prepared by the trial supervisor. They were stratified by sex and organised in blocks of 12. The randomisation lots were kept in envelopes, and the mother or guardian were asked to draw an envelope with an allocation lot from a bag.

No control vaccine or placebo was used. Both groups received vaccines. The allocation was not blinded as vaccines were registered on the child’s vaccination card, and it was indicated to health care staff not to give a missing DTP4 vaccination.

### Intervention

The DTP vaccine used in Guinea-Bissau was diphtheria-tetanus-whole-cell-pertussis from the Serum Institute of India from October 2005 to November 2007, and from Bio Pharma in Indonesia from November 2007 to end of trial. In September 2008, Guinea-Bissau started using pentavalent vaccine rather than DTP vaccine. At the same time, the policy of providing a booster dose was officially discontinued. The present trial continued the randomisation to DTP4+OPV4 vs OPV4-only, as per the approved protocol.

### Main Outcomes

The outcomes were all-cause non-accidental mortality and hospitalisation, as well as sex-difference in these outcomes.

### Follow-up and Assessment of outcomes

The children were followed from the date of inclusion to date of death, migration or 4 years of age, whichever came first. The trial team visited the children yearly after enrolment and finally when the children reached 4 years of age. The children taking part in the trial were also followed by the HDSS every 3 months.

Hospitalisation outcomes were followed through the BHP surveillance system at the paediatric ward of the nearby national hospital Simao Mendes, where most hospitalisations took place. Major diagnostic categories are registered for all hospital-admissions. Information on BHP trial numbers and ID were used to link the trial children to the hospital records. Additional fuzzy merge was performed using personal information (address, date of birth, name, mother’s name and sex) for hospital records with missing trial number or ID.

Deaths were detected both through the HDSS and through the surveillance at the paediatric ward. When a death was detected, a Guinean physician conducted a verbal autopsy with relatives.

### Sample size

With an expected 3% annual mortality rate and a 7% annual hospitalisation rate in the relevant age group, and with an expected total of 7,500 person-years of follow-up, we would be able to document a 35% reduction in mortality and a 23% reduction in hospitalisations.

### Other interventions and interactions

As the number of routine vaccinations and national health campaigns vaccinations increased through the 1990s and the 2000s, it has become increasingly clear that there are numerous interactions between different health interventions, such as vaccines and micronutrient supplementation, which are usually not taken into consideration in planning a vaccination programme. For example, the sequence of vaccinations, the time difference between non-live and live vaccines, and booster exposure to the same vaccines all had impact on the mortality levels. In addition, most vaccines have sex-differential NSEs (16).

Since children were enrolled at 18 months of age, there were numerous possibilities for interactions with (a) national health intervention campaigns before enrolment; (b) participation in previous RCTs; and (c) national health campaigns after enrolment in the trial.

### Statistical analysis

All children with follow-up and who received the per-protocol intervention were included in all analyses. Background factors are presented as percentages for categorical variables, while continuous variables are presented as mean with standard deviation by randomisation group. Assessment of differences in proportion among the randomisation groups were carried out using chi-squared or Fisher’s exact test. Distributions of continuous variables were compared using the Kruskal-Wallis rank-sum test. Deaths and observation time are presented with hazard ratios (HR) and Wald 95% confidence intervals (95% CI) estimated from a Cox proportional hazards model, with age as underlying time scale and with stratification by sex. Hospitalisations were analysed in recurrent-event Andersen-Gill Cox proportional hazards model including a 14-day grace period from date of discharge from the hospital. Age was inherently adjusted for in all Cox models. The proportional hazards assumption was assessed graphically and tested using Schoenfeld residuals.

According to the protocol adjustment for backgrounds factors should only be done for factors that changed the main results by 10%. None of the background factors fulfilled that criterion (data not shown). No imputation for missing data was done and no correction for multiple testing was applied.

### Ethical considerations

The study was explained to mothers/guardians of potential participants: “Though DTP is highly protective against whooping cough, it can occasionally give adverse reactions or limit the effect of measles vaccine. Your child has already received three doses of DTP. We would like to examine whether it is better to give just three doses of DTP or whether we should continue to give four doses. Thus, some children will receive DTP and OPV as is current practice and others will receive only OPV.” There was no patient or public involvement in the design, conduct and reporting of the trial. The children taking part in the study as well as those who refused to participate would benefit from additional follow up and free consultations.

The National Ethical Committee in Guinea-Bissau approved the protocol, and the Danish Central Ethical Committee gave consultative approval. The trial was registered on October 25^th^ 2005 at clinicaltrials.gov: https://clinicaltrials.gov/study/NCT00244673.

## RESULTS

Between October 2005 and October 2009, 5,960 children were enrolled in the DTP after MV part of the trial (arms 2 and 4, Supplementary Figure 1). Of the 114 children in arm 2 who did not receive the booster vaccination, 42 had received DTP+OPV and three died, whereas 72 received only OPV and three died. Of the 5,846 enrolled in the booster part of the trial (arm 4, Supplementary Figure 1) 82 were excluded due various errors, e.g. missing ID, duplicate entry, being less than 17 months of age, and not having received MV (Figure 1). Additionally, 90 children had no follow-up after enrolment. The analyses therefore included 5,674 children (OPV4+DTP4=2,864, OPV4=2,810).

**Figure 1.**
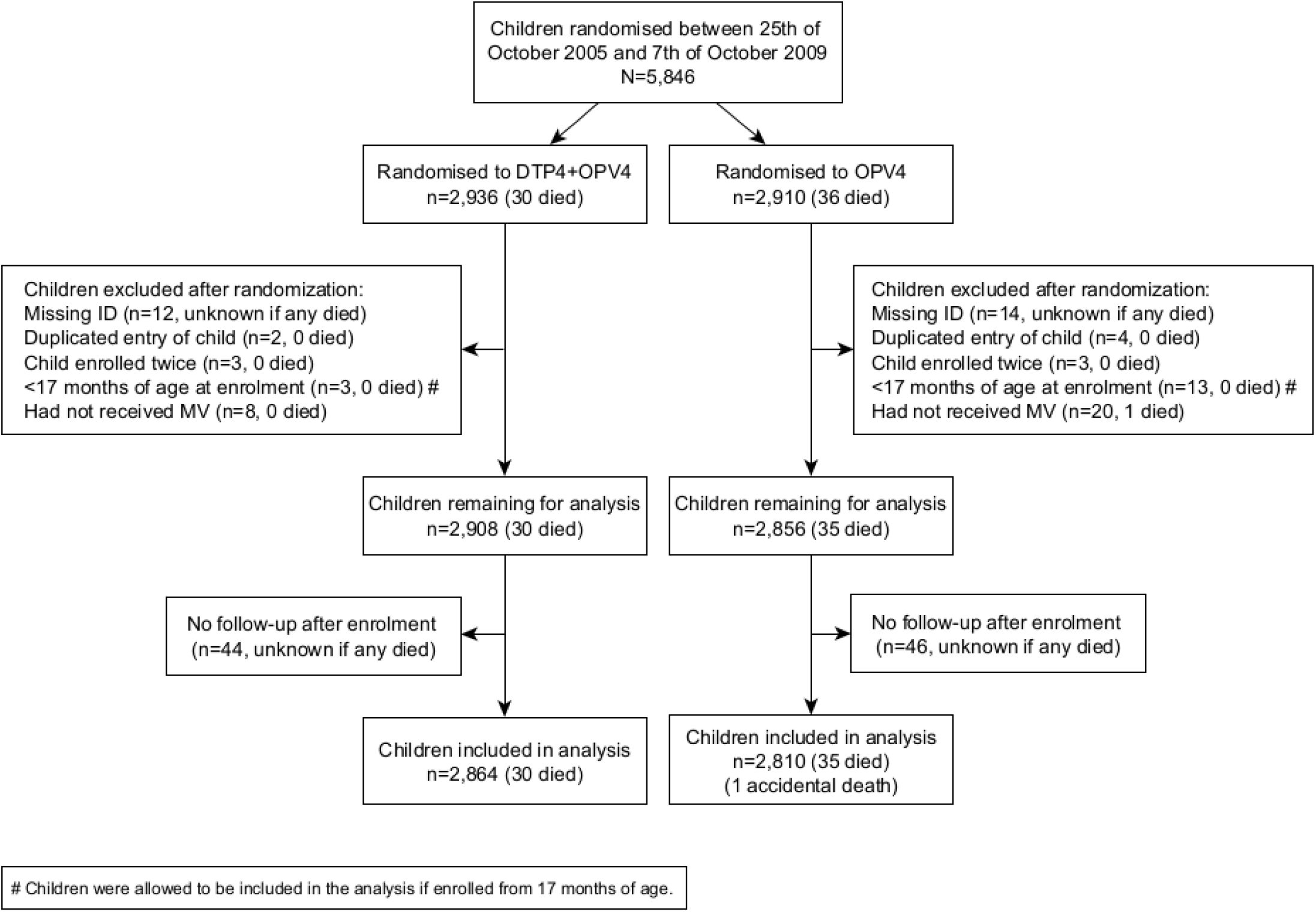
Trial flowchart.

The two randomisation groups were generally well balanced with respect to most background factors. However, the OPV4-only group tended to have had more hospitalisations prior to the enrolment and they had lower weight and height (Table 1).

**Table 1.** Baseline characteristics by randomisation group (DTP4+OPV4 and OPV4). Overall and by sex.

|  | **All** |  | **Males** |  | **Females** |  |
| --- | --- | --- | --- | --- | --- | --- |
|  | **DTP booster+OPV** | **OPV only** | **DTP booster+OPV** | **OPV only** | **DTP booster+OPV** | **OPV only** |
|  | **% (n)**<br>**[n missing]** | **% (n)**<br>**[n missing]** | **% (n)**<br>**[n missing]** | **% (n)**<br>**[n missing]** | **% (n)**<br>**[n missing]** | **% (n)**<br>**[n missing]** |
| Number of children | 2,864 | 2,810 | 1,433 | 1,403 | 1,431 | 1,407 |
| **Demographic factors** |  |  |  |  |  |  |
| Male sex | 50 (1433) [0] | 50 (1403) [0] | N/A | N/A | N/A | N/A |
| **Inclusion health center** |  |  |  |  |  |  |
| Bandim | 41 (1179) [0] | 42 (1168) [0] | 42 (609) [0] | 42 (587) [0] | 40 (570) [0] | 41 (581) [0] |
| Belem | 18 (520) [0] | 17 (490) [0] | 19 (270) [0] | 17 (239) [0] | 17 (250) [0] | 18 (251) [0] |
| Cuntum | 41 (1165) [0] | 41 (1152) [0] | 39 (554) [0] | 41 (577) [0] | 43 (611) [0] | 41 (575) [0] |
| **Inclusion time** |  |  |  |  |  |  |
| Enrolled in the morning | 24 (688) [0] | 23 (658) [0] | 24 (342) [0] | 23 (318) [0] | 24 (346) [0] | 24 (340) [0] |
| Enrolled in the afternoon | 76 (2176) [0] | 77 (2152) [0] | 76 (1091) [0] | 77 (1085) [0] | 76 (1085) [0] | 76 (1067) [0] |
| During the rainy season (June-Nov) | 48 (1373) [0] | 48 (1359) [0] | 48 (682) [0] | 48 (667) [0] | 48 (691) [0] | 49 (692) [0] |
| **Ethnicity** |  |  |  |  |  |  |
| Pepel | 30 (861) [14] | 31 (861) [9] | 30 (426) [9] | 30 (420) [6] | 30 (435) [5] | 31 (441) [3] |
| Other | 69 (1989) [14] | 69 (1940) [9] | 70 (998) [9] | 70 (977) [6] | 69 (991) [5] | 68 (963) [3] |
| **Risk factors** |  |  |  |  |  |  |
| Not breastfeeding | 62 (1769) [53] | 62 (1752) [44] | 61 (867) [22] | 62 (872) [28] | 63 (902) [31] | 63 (880) [16] |
| Previously hospitalized | 11 (329) [30] | 13 (369) [33] | 12 (176) [15] | 15 (206) [17] | 11 (153) [15] | 12 (163) [16] |
| Twin | 3 (96) [1] | 4 (103) [0] | 3 (43) [1] | 4 (55) [0] | 4 (53) [0] | 3 (48) [0] |
| Mother has died | 0 (3) [2] | 0 (3) [1] | 0 (1) [1] | 0 (3) [1] | 0 (2) [1] | 0 (0) [0] |
| Received medicine on the day of inclusion | 8 (230) [16] | 7 (204) [11] | 9 (128) [6] | 7 (100) [8] | 7 (102) [10] | 7 (104) [3] |
| **Symptoms on the day of inclusion** |  |  |  |  |  |  |
| Cough | 17 (494) [5] | 18 (519) [5] | 18 (264) [3] | 19 (264) [3] | 16 (230) [2] | 18 (255) [2] |
| Vomit | 0 (9) [3] | 1 (19) [4] | **0 (3) [2] #** | **1 (11) [3] #** | 0 (6) [1] | 1 (8) [1] |
| Diarrhoea | 7 (204) [3] | 7 (195) [4] | 8 (119) [2] | 8 (117) [4] | 6 (85) [1] | 6 (78) [0] |
| Respiratory count >40 cpm | 5 (148) [1181] | 5 (153) [1140] | 6 (86) [597] | 6 (82) [573] | 4 (62) [584] | 5 (71) [567] |
| Temperature >37 C | 2 (49) [2] | 2 (48) [1] | 2 (27) [1] | 2 (25) [0] | 2 (22) [1] | 2 (23) [1] |
| **Continuous background factors** |  |  |  |  |  |  |
| Inclusion age in months, mean (sd) | 20.0 (3.3) [0] | 19.8 (3.1) [0] | 19.9 (3.2) [0] | 19.8 (3.1) [0] | 20.0 (3.3) [0] | 19.8 (3.2) [0] |
| Mother's MUAC in mm, mean (sd) | 275.7 (37.5) [443] | 274.6 (37.9) [409] | 276.6 (37.9) [208] | 274.2 (36.3) [213] | 274.8 (37.1) [235] | 274.9 (39.4) [196] |
| Child weight in kg, mean (sd) | **10.4 (1.4) [1] #** | **10.3 (1.4) [2] #** | 10.6 (1.4) [0] | 10.5 (1.4) [0] | **10.1 (1.4) [1] #** | **10.0 (1.3) [2] #** |
| Child height in cm, mean (sd) | **81.3 (4.1) [8] #** | **80.9 (4.2) [3] #** | 81.8 (4.1) [4] | 81.5 (4.4) [2] | **80.8 (4.0) [4] #** | **80.4 (4.0) [1] #** |
| Child MUAC at enrolment in mm, mean (sd) | 146.9 (12.5) [1] | 146.5 (11.8) [0] | 147.7 (12.7) [0] | 147.5 (11.9) [0] | 146.1 (12.3) [1] | 145.5 (11.6) [0] |
| Child temperature in °C, mean (sd) | 36.1 (0.5) [2] | 36.1 (0.5) [1] | 36.1 (0.5) [1] | 36.1 (0.5) [0] | 36.1 (0.5) [1] | 36.1 (0.6) [1] |
| **Participation in other RCTs** |  |  |  |  |  |  |
| Enrolled in early MV trial (DDNOVO) | 58 (1661) [0] | 58 (1624) [0] | 58 (833) [0] | 58 (809) [0] | 58 (828) [0] | 58 (815) [0] |
| Received trial early MV (4-5 m) | 19 (544) [1203] | 19 (541) [1186] | 20 (288) [600] | 19 (262) [594] | 18 (256) [603] | 20 (279) [592] |
| Received trial late MV (18 m) | 18 (518) [1203] | 17 (476) [1186] | 18 (265) [600] | 17 (238) [594] | 18 (253) [603] | 17 (238) [592] |
| Enrolled in neonatal VAS trial | 68 (1935) [0] | 67 (1880) [0] | 69 (987) [0] | 68 (958) [0] | 66 (948) [0] | 66 (922) [0] |
| Received trial neonatal VAS | 42 (1192) [929] | 42 (1191) [930] | 42 (606) [446] | 43 (598) [445] | 41 (586) [483] | 42 (593) [485] |
| Enrolled in low birthweight BCG trial | 3 (74) [0] | 3 (92) [0] | 2 (35) [0] | 3 (42) [0] | 3 (39) [0] | 4 (50) [0] |
| Received trial BCG at birth | 1 (34) [2790] | 2 (43) [2718] | 1 (15) [1398] | 1 (18) [1361] | 1 (19) [1392] | 2 (25) [1357] |
| **Exposure to campaigns before enrolment (based on eligibility)** |  |  |  |  |  |  |
| OPV campaign | 57 (1619) [0] | 56 (1581) [0] | 55 (788) [0] | 55 (771) [0] | 58 (831) [0] | 58 (810) [0] |
| OPV+VAS campaign | 41 (1169) [0] | 40 (1122) [0] | 39 (565) [0] | 38 (534) [0] | 42 (604) [0] | 42 (588) [0] |
| VAS campaign | 78 (2244) [0] | 80 (2235) [0] | 79 (1138) [0] | 80 (1127) [0] | 77 (1106) [0] | 79 (1108) [0] |
| MV+VAS campaign | 13 (359) [0] | 12 (340) [0] | 12 (179) [0] | 12 (172) [0] | 13 (180) [0] | 12 (168) [0] |

### Mortality and morbidity

The was no difference in non-accidental mortality with 30 and 35 deaths in the two randomisation groups, the HR for DTP4+OPV4 vs OPV4 being 0.84 (0.52-1.37)(Table 2). The mortality rate in the trial was 0.55% (65/11,813 person-years(PYRS)). Since the trial was planned with a mortality rate of 3%, the observed rate was 82% lower than expected.

**Table 2.** Rates of non-accidental deaths, hospitalisations overall and by major categories per 100 person-years overall, by randomisation group and sex. Cox regression analysis yielding HRs of DTP4+OPV4 vs. OPV overall and by sex. Follow-up from enrolment to 4 years of age.

|  | Hospitalisation rate per 100 person-years (hospitalisations/person-years) |  |  |  |  |  | HR of DTP4+OPV4 vs OPV4 (95% CI) |  |  |
| --- | --- | --- | --- | --- | --- | --- | --- | --- | --- |
|  | DTP4+OPV4 |  |  | OPV4 |  |  |  |  |  |
|  | All | Males | Females | All | Males | Females | All | Males | Females |
| Non-accidental mortality | 0.50<br>(30/5971) | 0.47<br>(14/2981) | 0.54<br>(16/2990) | 0.60<br>(35/5841) | 0.55<br>(16/2930) | 0.65<br>(19/2911) | 0.84 (0.52-1.37) | 0.86 (0.42-1.76) | 0.82 (0.42-1.60) |
| All-cause hospitalisations | 4.60<br>(274/5962) | 4.97<br>(148/2976) | 4.22<br>(126/2986) | 5.08<br>(296/5831) | 5.71<br>(167/2925) | 4.44<br>(129/2907) | 0.91 (0.77-1.07) | 0.87 (0.70-1.09) | 0.96 (0.75-1.22) |
| By hospitalisation cause #1 |  |  |  |  |  |  |  |  |  |
| Malaria | 2.75<br>(164/5962) | 3.09<br>(92/2976) | 2.41<br>(72/2986) | 3.26<br>(190/5831) | 3.52<br>(103/2925) | 2.99<br>(87/2907) | 0.85 (0.69-1.04) | 0.88 (0.66-1.16) | 0.81 (0.59-1.10) |
| Respiratory infection | 1.11<br>(66/5962) | 1.08<br>(32/2976) | 1.14<br>(34/2986) | 1.18<br>(69/5831) | 1.54<br>(45/2925) | 0.83<br>(24/2907) | 0.94 (0.67-1.32) | 0.70 (0.44-1.10) | 1.39 (0.83-2.35) |
| Gastrointestinal infection | 0.34<br>(20/5962) | 0.20<br>(6/2976) | 0.47<br>(14/2986) | 0.31<br>(18/5831) | 0.27<br>(8/2925) | 0.34<br>(10/2907) | 1.10 (0.58-2.07) | 0.74 (0.26-2.13) | 1.38 (0.61-3.11) |
| Other infection | 0.15<br>(9/5962) | 0.17<br>(5/2976) | 0.13<br>(4/2986) | 0.21<br>(12/5831) | 0.27<br>(8/2925) | 0.14<br>(4/2907) | 0.74 (0.31-1.75) | 0.61 (0.20-1.88) | 0.98 (0.25-3.93) |
| Other causes | 0.23<br>(14/5962) | 0.40<br>(12/2976) | 0.07<br>(2/2986) | 0.12<br>(7/5831) | 0.10<br>(3/2925) | 0.14<br>(4/2907) | 1.96 (0.79-4.85) | 3.92 (1.11-13.9) | 0.49 (0.09-2.67) |

#1 One hospitalisation excluded with missing cause.

There were 571 hospitalisations in the study cohort, with 274 and 297 in each group. The hazard ratio was 0.91 (0.77-1.07)(Table 2).

There were no differences in F/M mortality ratios (Supplementary Table 2). However, there were significantly lower F/M hospitalisation ratios in the OPV4-only group, particularly for respiratory infectious hospitalisations (Supplementary Table 2).

### Interactions

Since it has become increasingly clear that health interventions interact and many interventions were introduced in the trial period, possible interactions were explored, even though it was clear that the power was low. Firstly, with campaigns and RCT interventions administered before the children were enrolled in the DTP4-trial. Secondly, with campaigns administered after enrolment in the trial (Supplementary Tables 3 and 4).

All children had been eligible to receive at least one health intervention campaign before enrolment in the DTP4 trial (Table 1). Furthermore, 77% of trial participants had previously taken part in another randomised trial (Supplementary Table 5). In addition, the vast majority of children were exposed to further national campaigns with OPV-only, OPV+vitamin A supplementation (VAS), VAS-only, MV+VAS, or H1N1 influenza vaccine during the conduct of the trial (Figure 2). For only 12% of the follow-up time (and 17% of deaths) had the trial participants not received another campaign after enrolment.

**Figure 2.**
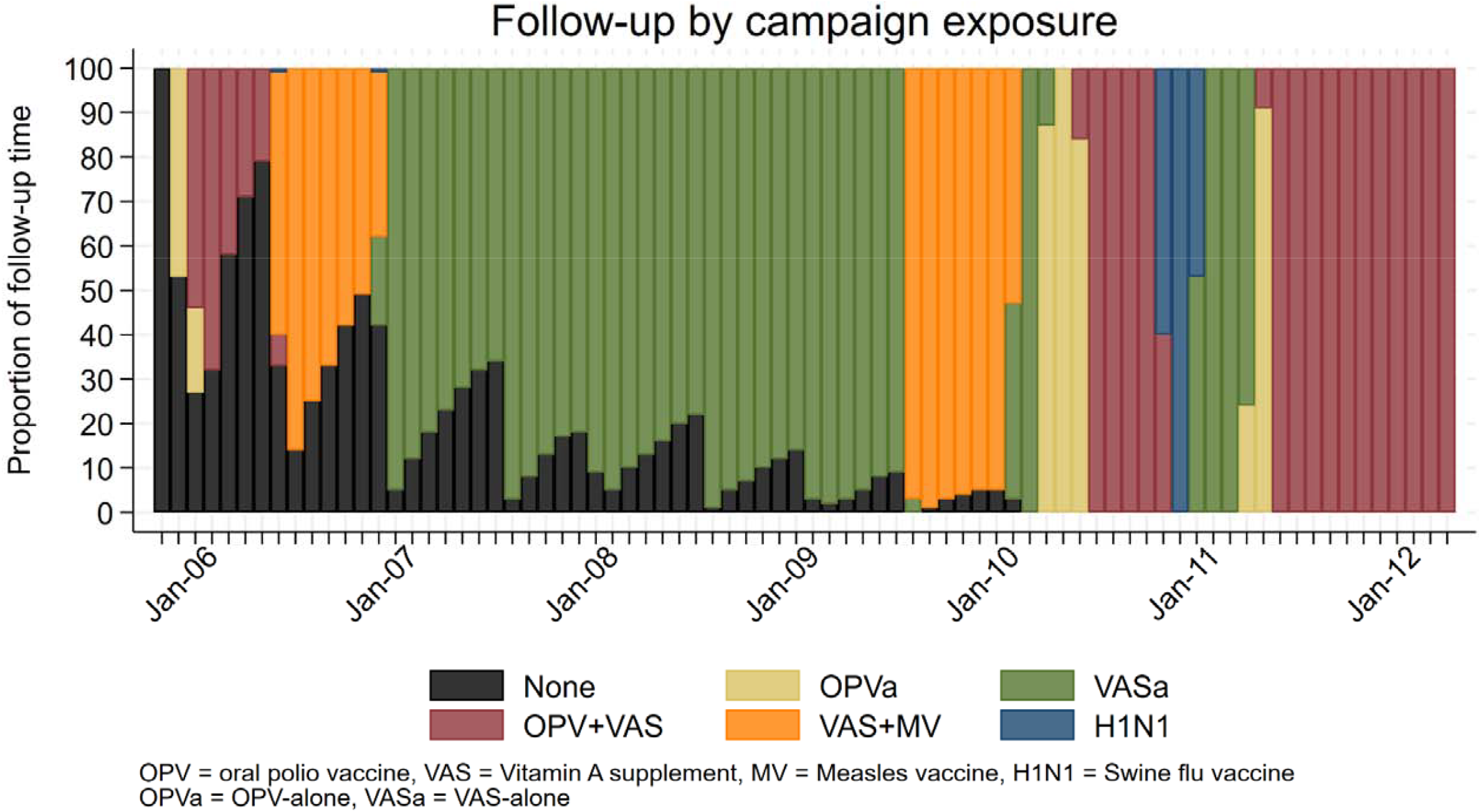
Proportion of exposure to different campaigns during follow-up.

There was one significant interaction (Supplementary Table 3): between the periods where Serum Institute of India (SII) DTP (MR=0.54 (0.27-1.06)) and DTP from Bio Pharma, Indonesia (MR=1.48 (0.69-3.15)) was used (test of interaction, p=0.05), this was observed only for females (test of interaction, p=0.04; data not shown)

## DISCUSSION

The study hypotheses were not supported. Randomisation to DTP4+OPV4 was not associated with higher mortality rates or higher hospital admission rates nor were there major sex differences. There were significantly fewer females than male admissions in the OPV4 group, as hypothesised, but it was not significantly different from the pattern in the DTP4+OPV4 group (Supplementary Table 4).

The RCT was strongly underpowered. After the civil war in 1998-1999 and the following disruption of the health care system, partly due to health care workers fleeing Guinea-Bissau during the war, and a measles epidemic in 2003, the expectation was an annual mortality rate of 3%. Fortunately for the children, the rate declined to 0.55% in the trial period 2005-2012.

### Consistency with previous studies

To our knowledge, there has been no previous RCT of DTP4 vs no DTP4 examining the effect on mortality. In a previous trial, we used a booster dose of BCG at 19 months of age to modify the expected negative effect of DTP4 assumed to be given at 18 months. If BCG was given after DTP4, there was a 64% (1-87%) reduction in mortality up to 5 years of age. However, if DTP4 was not given before enrolment at 19 months and DTP4 therefore was likely to be given after BCG, mortality was increased (mortality ratio 78% (4-204%))(17).

The are several observational studies of DTP4 which have pointed to increased female mortality after DTP4 (9-14,18). The present RCT is therefore an outlier which needs an explanation. The drop in power due to the declining mortality rate may not only have lowered the possibility of finding significant tendencies; it is also likely that the immune mediated NSEs are more pronounced when mortality is high, so when mortality declines by >80%, the residual deaths may be less likely to be affected by immunological changes.

Initially, trials of NSEs were planned more or less as vaccine efficacy studies. However, it has become increasingly clear that there are interactions with other routine vaccinations, vaccination campaigns, and other interventions affecting the immune system like vitamin A (16,19,20). Hence, in the present RCT we examined possible interactions with campaigns before enrolment, previous RCTs, and campaigns given after enrolment. There was limited power in the study and too many possible interactions so there was no definitive pattern, except that the two different strains of DTP had different effects; however, this could be due to other temporal factors, such as the campaigns conducted in the respective periods (Figure 2) rather than inherent effects of the strains of DTP.

## CONCLUSION

In conclusion, the trial did not find the expected beneficial effect of not given DTP4. From the point of view of a planned RCT, this trial failed in several respects. It was strongly underpowered because child mortality declined much more than expected. Due to the numerous campaigns a limited part of the follow-up time was a comparison between DTP4+OPV4 vs OPV4 as the most recently received vaccinations.

### Postscript

We apologise for the late reporting. The implementation of the trial went quite different from the scheduled plans. In this older age group, more children than expected were registered by an ID and address that could not be followed. Funding was lacking for the PhD student to complete the data cleaning and analysis. Before funding could be obtained, the Guinean field supervisor had died which made it difficult to resolve some inconsistencies in data. The senior authors had too many other commitments. Finally, from 2020, the COVID-19 pandemic changed all priorities.

## Supporting information

Main text DTP..maintext,docx

## Contributors

PA wrote the first draft of the paper. The corresponding author attests that all listed authors meet authorship criteria and that no others meeting the criteria have been omitted.

## Transparency

The senior authors affirms that the manuscript is an honest, accurate, and transparent account of the study being reported and that no important aspects of the study have been omitted.

## Data availability statement

By contact to the corresponding author.

