## Supplementary material for "Randomised trial of not providing booster diphtheria-tetanus-pertussis (DTP) vaccination after measles vaccination and child survival: A failed trial": Main text DTP..maintext,docx

**Supplementary Figure 1**. Trial study design for children to receive *either* DTP with MV *or* DTP after MV. Arm 4 is presented in the main text.

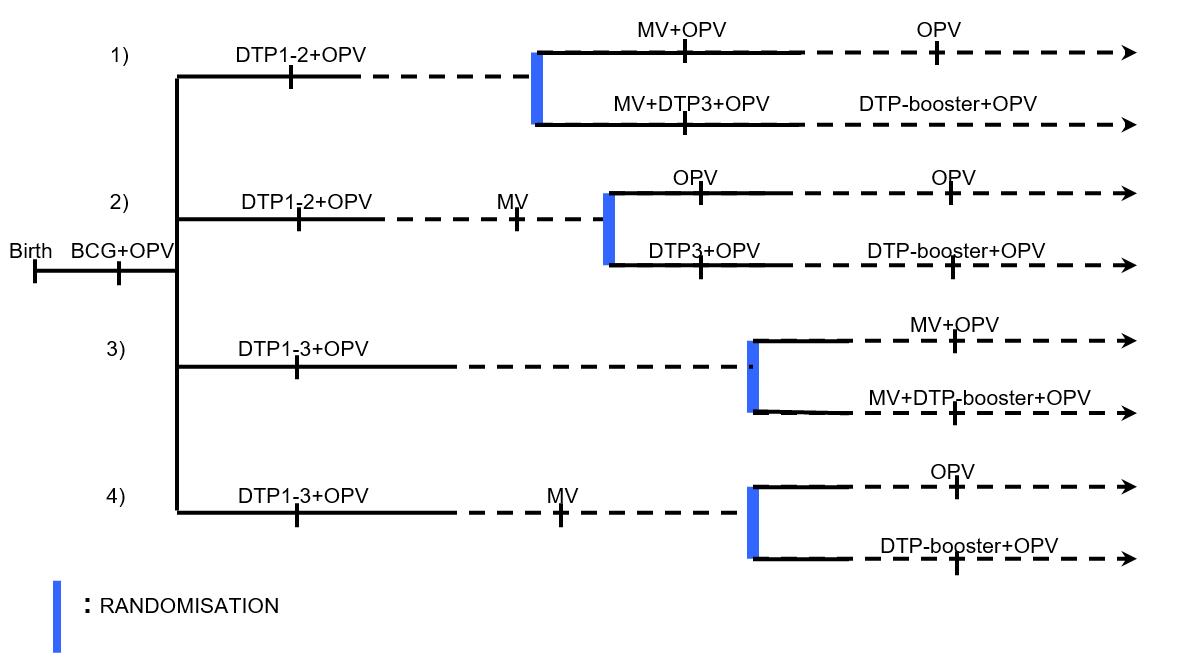

**Supplementary Table 1. Mortality ratio for children receiving DTP with MV (DTP=MV) or after MV (DTP>MV) compared with children having received MV as last vaccination (DTP<MV)**

| **A. MV and DTP administered simultaneously (MV=DTP)** | | |  |
| --- | --- | --- | --- |
| **Country** | **Number of children** | **Study** | **MR for MV=DTP**  **vs DTP<MV** |
| Congo (9) |  | EZ-trial at 6 months; followed to 36 months | 5.38 (1.37-21.2) |
| Malawi (10) | 751 | Routine vaccination | 5.27 (1.11-25.0) |
| Senegal (11) | 4,135 | Routine vaccination – 9-24 months: 1996-99 | 2.21 (1.10-4.43) |
| Guinea-Bissau (12) | 403 | Hospital case fatality 6-17 months; 1990-96 and 2001 | 1.87 (1.06-3.31) |
| Ghana (13) | 6,761 | Routine vaccination; mortality within 12 months | 1.76 (1.05-2.94) |
| **Vaccination coverage for dead children versus community controls** | | | **Relative risk** |
| Gambia (14) |  | 22% (9/41) of dead children aged 9-17 months had received DTP>=MV compared with 6% (163/2539) in community survey | 3.42 (1.88-6.21) |
| **Combined estimate** | | | 2.33 (1.76-3.09) |
| **B. DTP administered after MV (DTP>MV)** | | |  |
| **Country** | **Number of children** | **Study** | **MR for DTP>MV**  **vs DTP<MV** |
| Senegal (11) | 4135 | Routine vaccination – 9-24 months: 1996-99 | 2.72 (1.16-6.39) |
| Guinea-Bissau (12) | 276 | Hospital case fatality 6-17 months; 1990-96 and 2001 | 1.47 (0.68-3.20) |
| Ghana (13) | 6,761 | Routine vaccination; mortality within 12 months | 1.68 (0.94-2.99) |
| **Combined estimate** | | | 1.81 (1.20-2.72) |

**Supplementary Table 2. Female-male HRs of non-accidental mortality and hospitalisations. Follow-up from enrolment to 4 years of age.**

|  | **HR of females vs males (95% CI)** | | |
| --- | --- | --- | --- |
|  | **All** | **DTP4+OPV4** | **OPV4** |
| Non-accidental mortality | 1.16 (0.71-1.89) | 1.14 (0.56-2.34) | 1.17 (0.60-2.28) |
| All-cause hospitalisations | 0.81 (0.69-0.96) | 0.85 (0.67-1.08) | 0.78 (0.62-0.98) |
| **By hospitalisation cause #1** | | | |
| Malaria | 0.82 (0.66-1.01) | 0.78 (0.57-1.06) | 0.85 (0.64-1.13) |
| Respiratory infection | 0.76 (0.54-1.06) | 1.07 (0.66-1.73) | 0.53 (0.33-0.88) |
| Gastrointestinal infection | 1.72 (0.89-3.33) | 2.35 (0.90-6.11) | 1.26 (0.50-3.18) |
| Other infection | 0.62 (0.26-1.49) | 0.80 (0.22-2.99) | 0.50 (0.15-1.66) |
| Other causes | 0.40 (0.16-1.03) | 0.17 (0.04-0.74) | 1.33 (0.30-5.96) |

*#1 One hospitalisation excluded with missing cause.*

**Supplementary Table 3. Explorative analysis of the impact of contextual factors on the all-cause mortality hazard ratio (HR).**

| **Contextual factor** | **Mortality rate per 100 person-years (deaths/person-years)** | | **HR of DTP4+OPV4 vs OPV4 (95% CI)** | **Female/Male HR (95% CI)** |
| --- | --- | --- | --- | --- |
|  | **DTP4+OPV4** | **OPV4** |  |  |
| **Participation in other RCTs** | | | | |
| Received no trial MV | 0.59 (22/3715) | 0.66 (24/3619) | 0.89 (0.50-1.60) | 1.07 (0.60-1.92) |
| Received early (4m) MV in DDNOVO | 0.34 (4/1160) | 0.60 (7/1176) | 0.58 (0.17-1.99) | 1.23 (0.38-4.04) |
| Received late (18m) MV in DDNOVO | 0.36 (4/1098) | 0.38 (4/1046) | 0.97 (0.24-3.87) | 1.72 (0.41-7.22) |
| Received no trial NVAS | 0.67 (23/3437) | 0.51 (17/3309) | 1.30 (0.69-2.43) | 1.64 (0.86-3.11) |
| Received trial NVAS | 0.28 (7/2534) | 0.71 (18/2532) | 0.39 (0.16-0.94) | 0.68 (0.31-1.52) |
| **Campaigns after enrolment** | | | | |
| Not yet received any Campaign | 0.71 (5/707) | 0.86 (6/699) | 0.82 (0.25-2.68) | 0.58 (0.17-1.97) |
| Received any Campaign | 0.47 (25/5264) | 0.56 (29/5143) | 0.85 (0.50-1.44) | 1.34 (0.78-2.30) |
| Not yet received C-OPV | 0.48 (25/5262) | 0.66 (34/5190) | 0.72 (0.43-1.21) | 1.03 (0.62-1.72) |
| Received C-OPV | 0.70 (5/709) | 0.15 (1/652) | 4.51 (0.53-38.7) | 5.26 (0.61-45.0) |
| Not yet received C-VAS | 1.01 (9/890) | 1.25 (11/878) | 0.79 (0.33-1.90) | 0.97 (0.40-2.32) |
| Received C-VAS | 0.41 (21/5081) | 0.48 (24/4963) | 0.86 (0.48-1.54) | 1.25 (0.70-2.26) |
| Not yet received C-OPV+VAS | 0.50 (27/5359) | 0.66 (35/5279) | 0.76 (0.46-1.25) | 1.14 (0.69-1.87) |
| Received C-OPV+VAS | 0.49 (3/612) | 0.00 (0/562) | - | 2.08 (0.19-22.9) |
| Not yet received C-MV+VAS | 0.45 (22/4891) | 0.58 (28/4813) | 0.77 (0.44-1.35) | 0.93 (0.53-1.62) |
| Received C-MV+VAS | 0.74 (8/1080) | 0.68 (7/1028) | 1.11 (0.40-3.07) | 2.57 (0.82-8.07) |
| Not yet received C-H1N1 | 0.51 (29/5737) | 0.62 (35/5634) | 0.81 (0.50-1.33) | 1.13 (0.69-1.85) |
| Received C-H1N1 | 0.43 (1/234) | 0.00 (0/207) | - | - |
| **Campaigns before enrolment** | | | | |
| Not received any campaign | 0 (0/0) | 0 (0/0) | - | - |
| Received any campaign | 0.50 (30/5971) | 0.60 (35/5841) | 0.84 (0.52-1.37) | 1.17 (0.72-1.90) |
| Not C-OPV | 0.72 (19/2640) | 0.59 (15/2553) | 1.23 (0.63-2.42) | 1.19 (0.61-2.34) |
| Received C-OPV | 0.33 (11/3331) | 0.61 (20/3289) | 0.54 (0.26-1.13) | 1.16 (0.57-2.36) |
| Not received C-OPV+VAS | 0.61 (22/3600) | 0.57 (20/3528) | 1.08 (0.59-1.98) | 1.05 (0.57-1.92) |
| Received C-OPV+VAS | 0.34 (8/2371) | 0.65 (15/2313) | 0.52 (0.22-1.23) | 1.44 (0.63-3.34) |
| Not received C-VAS | 0.31 (4/1285) | 0.67 (8/1195) | 0.46 (0.14-1.54) | 1.82 (0.55-6.06) |
| Received C-VAS | 0.55 (26/4686) | 0.58 (27/4646) | 0.96 (0.56-1.64) | 1.06 (0.62-1.82) |
| Not received C-MV+VAS | 0.44 (23/5260) | 0.60 (31/5184) | 0.73 (0.43-1.26) | 1.25 (0.73-2.14) |
| Received C-MV+VAS | 0.98 (7/711) | 0.61 (4/657) | 1.62 (0.47-5.54) | 0.83 (0.25-2.72) |
| **Other temporal factors** | | | | |
| DTP vaccine (Serum Institute of India) | 0.34 (13/3866) | 0.63 (24/3828) | 0.54 (0.27-1.06) | 0.93 (0.49-1.77) |
| DTP vaccine (Bio Pharma) | 0.81 (17/2105) | 0.55 (11/2014) | 1.48 (0.69-3.15) | 1.61 (0.75-3.43) |
| Enrolled in rainy season | 0.49 (14/2877) | 0.57 (16/2825) | 0.86 (0.42-1.77) | 0.98 (0.48-2.00) |
| Enrolled in dry season | 0.52 (16/3094) | 0.63 (19/3016) | 0.82 (0.42-1.59) | 1.37 (0.70-2.68) |
| Follow-up in rainy season | 0.46 (24/5271) | 0.52 (27/5161) | 0.87 (0.50-1.52) | 1.13 (0.65-1.95) |
| Follow-up in dry season | 0.11 (6/5243) | 0.16 (8/5113) | 0.73 (0.25-2.12) | 1.34 (0.47-3.87) |
| 18-23 months of age | 0.95 (10/1053) | 1.14 (12/1053) | 0.82 (0.36-1.91) | 0.57 (0.24-1.37) |
| 24-35 months of age | 0.47 (12/2552) | 0.68 (17/2493) | 0.69 (0.33-1.45) | 1.90 (0.88-4.08) |
| 36-47 months of age | 0.34 (8/2365) | 0.26 (6/2296) | 1.29 (0.45-3.72) | 1.33 (0.46-3.84) |
| Follow-up Calendar year 2005-2006 | 0.34 (2/587) | 1.06 (6/564) | 0.31 (0.06-1.56) | 1.56 (0.37-6.53) |
| Follow-up Calendar year 2007 | 0.56 (8/1417) | 0.86 (12/1394) | 0.68 (0.28-1.65) | 0.64 (0.26-1.58) |
| Follow-up Calendar year 2008 | 0.50 (9/1810) | 0.61 (11/1808) | 0.82 (0.34-1.97) | 1.53 (0.63-3.75) |
| Follow-up Calendar year 2009 | 0.37 (5/1363) | 0.37 (5/1336) | 0.97 (0.28-3.35) | 0.70 (0.20-2.48) |
| Follow-up Calendar year 2010-2012 | 0.76 (6/794) | 0.14 (1/740) | 5.48 (0.66-45.6) | 6.41 (0.77-53.2) |
| Inclusion year 2005 | 0.00 (0/198) | 0.55 (1/183) | - | - |
| Inclusion year 2006 | 0.34 (7/2052) | 0.64 (13/2026) | 0.53 (0.21-1.33) | 1.37 (0.56-3.34) |
| Inclusion year 2007 | 0.43 (8/1844) | 0.59 (11/1857) | 0.74 (0.30-1.85) | 0.62 (0.25-1.58) |
| Inclusion year 2008 | 0.69 (10/1447) | 0.58 (8/1390) | 1.19 (0.47-3.01) | 1.65 (0.64-4.25) |
| Inclusion year 2009 | 1.16 (5/431) | 0.52 (2/385) | 2.27 (0.44-11.7) | 1.40 (0.31-6.24) |
| Birth year 2001-2004 | 0.21 (3/1452) | 0.58 (8/1389) | 0.36 (0.10-1.36) | 1.60 (0.47-5.48) |
| Birth year 2005 | 0.41 (8/1964) | 0.70 (14/2001) | 0.58 (0.24-1.39) | 0.82 (0.35-1.89) |
| Birth year 2006 | 0.59 (10/1694) | 0.60 (10/1673) | 0.99 (0.41-2.38) | 1.29 (0.54-3.12) |
| Birth year 2007-2008 | 1.04 (9/861) | 0.39 (3/778) | 2.72 (0.74-10.0) | 1.55 (0.49-4.88) |

*OPV=oral polio vaccine, VAS=vitamin A supplementation, MV=measles vaccine, H1N1=swine flu vaccine.*
**Supplementary Table 4. Explorative analysis of the impact of contextual factors on the hospitalisation hazard ratio (HR).**

| **Contextual factor** | **Mortality rate per 100 person-years (deaths/person-years)** | | **HR of DTP4+OPV4 vs OPV4 (95% CI)** | **Female/Male HR (95% CI)** |
| --- | --- | --- | --- | --- |
|  | **DTP4+OPV4** | **OPV4** |  |  |
| **Participation in other RCTs** | | | | |
| Received no trial MV | 4.85 (180/3709) | 5.09 (184/3613) | 0.96 (0.78-1.18) | 0.76 (0.61-0.93) |
| Received early (4m) MV in DDNOVO | 3.88 (45/1159) | 5.54 (65/1174) | 0.70 (0.48-1.02) | 0.89 (0.61-1.29) |
| Received late (18m) MV in DDNOVO | 4.47 (49/1097) | 4.50 (47/1044) | 1.00 (0.67-1.49) | 0.95 (0.64-1.42) |
| Received no trial NVAS | 4.81 (165/3431) | 5.45 (180/3303) | 0.88 (0.71-1.09) | 0.73 (0.59-0.90) |
| Received trial NVAS | 4.31 (109/2531) | 4.59 (116/2528) | 0.94 (0.73-1.23) | 0.95 (0.73-1.23) |
| **Campaigns after enrolment** | | | | |
| Not yet received any Campaign | 6.94 (49/706) | 8.90 (62/696) | 0.78 (0.54-1.14) | 0.76 (0.52-1.11) |
| Received any Campaign | 4.28 (225/5256) | 4.56 (234/5135) | 0.94 (0.78-1.13) | 0.82 (0.69-0.99) |
| Not yet received C-OPV | 4.59 (241/5254) | 5.17 (268/5181) | 0.89 (0.75-1.06) | 0.82 (0.69-0.98) |
| Received C-OPV | 4.66 (33/708) | 4.30 (28/651) | 1.08 (0.65-1.78) | 0.73 (0.44-1.22) |
| Not yet received C-VAS | 7.10 (63/888) | 8.68 (76/876) | 0.82 (0.59-1.15) | 0.79 (0.56-1.10) |
| Received C-VAS | 4.16 (211/5074) | 4.44 (220/4956) | 0.94 (0.78-1.13) | 0.82 (0.68-0.99) |
| Not yet received C-OPV+VAS | 4.58 (245/5351) | 5.26 (277/5270) | 0.87 (0.73-1.03) | 0.79 (0.67-0.94) |
| Received C-OPV+VAS | 4.75 (29/611) | 3.38 (19/562) | 1.44 (0.81-2.57) | 1.02 (0.58-1.79) |
| Not yet received C-MV+VAS | 4.46 (218/4884) | 5.06 (243/4805) | 0.88 (0.74-1.06) | 0.83 (0.69-0.99) |
| Received C-MV+VAS | 5.19 (56/1079) | 5.16 (53/1026) | 1.01 (0.69-1.47) | 0.74 (0.50-1.08) |
| Not yet received C-H1N1 | 4.54 (260/5728) | 5.08 (286/5625) | 0.89 (0.76-1.06) | 0.82 (0.69-0.97) |
| Received C-H1N1 | 5.99 (14/234) | 4.83 (10/207) | 1.28 (0.57-2.89) | 0.66 (0.29-1.52) |
| **Campaigns before enrolment** | | | | |
| Not received any campaign | - | - | - | - |
| Received any campaign | 4.60 (274/5962) | 5.08 (296/5831) | 0.91 (0.77-1.07) | 0.81 (0.69-0.96) |
| Not C-OPV | 5.62 (148/2636) | 5.49 (140/2548) | 1.03 (0.81-1.29) | 0.85 (0.67-1.07) |
| Received C-OPV | 3.79 (126/3327) | 4.75 (156/3283) | 0.80 (0.63-1.01) | 0.78 (0.62-0.99) |
| Not received C-OPV+VAS | 5.40 (194/3593) | 5.57 (196/3522) | 0.97 (0.80-1.19) | 0.81 (0.67-0.99) |
| Received C-OPV+VAS | 3.38 (80/2369) | 4.33 (100/2310) | 0.78 (0.58-1.05) | 0.83 (0.62-1.11) |
| Not received C-VAS | 2.96 (38/1284) | 3.85 (46/1194) | 0.77 (0.50-1.19) | 0.79 (0.51-1.21) |
| Received C-VAS | 5.04 (236/4678) | 5.39 (250/4638) | 0.94 (0.79-1.12) | 0.82 (0.69-0.98) |
| Not received C-MV+VAS | 4.63 (243/5252) | 5.02 (260/5175) | 0.92 (0.78-1.10) | 0.82 (0.69-0.97) |
| Received C-MV+VAS | 4.36 (31/710) | 5.49 (36/656) | 0.80 (0.49-1.29) | 0.76 (0.47-1.24) |
| **Other temporal factors** | | | | |
| DTP vaccine (Serum Institute of India) | 3.91 (151/3861) | 4.95 (189/3821) | 0.79 (0.64-0.98) | 0.77 (0.62-0.95) |
| DTP vaccine (Bio Pharma) | 5.85 (123/2101) | 5.32 (107/2010) | 1.10 (0.85-1.43) | 0.89 (0.69-1.15) |
| Enrolled in rainy season | 4.59 (132/2873) | 5.60 (158/2820) | 0.82 (0.65-1.04) | 0.84 (0.67-1.06) |
| Enrolled in dry season | 4.60 (142/3089) | 4.58 (138/3011) | 1.01 (0.80-1.27) | 0.78 (0.62-0.99) |
| Follow-up in rainy season | 5.40 (163/3017) | 5.94 (175/2949) | 0.91 (0.74-1.13) | 0.85 (0.69-1.05) |
| Follow-up in dry season | 3.77 (111/2945) | 4.20 (121/2883) | 0.90 (0.70-1.17) | 0.76 (0.58-0.98) |
| 18-23 months of age | 6.47 (68/1051) | 8.96 (94/1049) | 0.72 (0.53-0.98) | 0.81 (0.59-1.10) |
| 24-35 months of age | 4.79 (122/2548) | 5.22 (130/2488) | 0.92 (0.72-1.17) | 0.83 (0.64-1.06) |
| 36-47 months of age | 3.56 (84/2363) | 3.14 (72/2294) | 1.13 (0.83-1.55) | 0.79 (0.58-1.09) |
| Follow-up Calendar year 2005-2006 | 5.63 (33/586) | 6.58 (37/562) | 0.88 (0.55-1.41) | 0.97 (0.61-1.55) |
| Follow-up Calendar year 2007 | 3.18 (45/1416) | 5.75 (80/1391) | 0.56 (0.39-0.81) | 0.77 (0.54-1.09) |
| Follow-up Calendar year 2008 | 5.04 (91/1807) | 4.82 (87/1805) | 1.05 (0.78-1.40) | 0.74 (0.55-1.00) |
| Follow-up Calendar year 2009 | 5.00 (68/1360) | 4.35 (58/1334) | 1.15 (0.81-1.63) | 0.89 (0.62-1.26) |
| Follow-up Calendar year 2010-2012 | 4.67 (37/793) | 4.60 (34/739) | 1.01 (0.64-1.61) | 0.82 (0.52-1.32) |
| Inclusion year 2005 | 3.54 (7/198) | 2.19 (4/183) | 1.80 (0.52-6.19) | 1.23 (0.37-4.02) |
| Inclusion year 2006 | 3.27 (67/2050) | 4.30 (87/2023) | 0.76 (0.55-1.05) | 0.84 (0.61-1.15) |
| Inclusion year 2007 | 4.51 (83/1841) | 5.50 (102/1854) | 0.82 (0.62-1.10) | 0.68 (0.51-0.92) |
| Inclusion year 2008 | 6.09 (88/1444) | 5.19 (72/1387) | 1.18 (0.86-1.61) | 1.05 (0.77-1.43) |
| Inclusion year 2009 | 6.75 (29/430) | 8.07 (31/384) | 0.85 (0.51-1.42) | 0.65 (0.38-1.09) |
| Birth year 2001-2004 | 3.10 (45/1450) | 3.61 (50/1387) | 0.87 (0.58-1.31) | 0.85 (0.57-1.28) |
| Birth year 2005 | 4.23 (83/1961) | 5.66 (113/1997) | 0.75 (0.57-1.00) | 0.71 (0.53-0.94) |
| Birth year 2006 | 5.20 (88/1691) | 4.73 (79/1671) | 1.10 (0.81-1.49) | 1.02 (0.75-1.38) |
| Birth year 2007-2008 | 6.75 (58/859) | 6.95 (54/777) | 0.99 (0.68-1.44) | 0.74 (0.51-1.08) |

*OPV=oral polio vaccine, VAS=vitamin A supplementation, MV=measles vaccine, H1N1=swine flu vaccine.*

**Supplementary table 5. Overview of participation in other clinical trials among children enrolled in the DTP trial.**

| **RCT (enrolment period in DTP RCT)** | **% enrolled among the 5,674 DTP RCT children (N)** | **Intervention** | **% enrolled of DTP children (N)** |
| --- | --- | --- | --- |
| 2-dose MV RCT (25th of October 2005 to 4th of September 2008) | 58% (3,285) | No intervention/placebo | 21% (1,207) |
|  |  | Early MV (4.5 months of age) | 19% (1,085)# |
|  |  | Late MV (18 months of age) | 18% (994)# |
| VAS at birth RCTs (25^th^ of October 2005 to 19^th^ of June 2009) | 67% (3,815) | No intervention/placebo | 25% (1,432) |
|  |  | VAS | 42% (2,383) |
| BCG at birth to low birthweight children (29^th^ of June 2006 to 22^nd^ of September 2009) | 2.9% (166) | No intervention/placebo | 1.6% (89) |
|  |  | BCG | 1.4% (77) |
| Participation in any of the trials listed above | 77% (4,386) | No intervention | 39% (2,231) |
|  |  | 1 intervention | 42% (2,357) |
|  |  | 2 interventions | 19% (1,076) |
|  |  | 3 interventions | 0.2% (10) |

*#One child had received both the early and late MV.*
